# Systematic review and patient-level meta-analysis of SARS-CoV-2 viral dynamics to model response to antiviral therapies

**DOI:** 10.1101/2020.08.20.20178699

**Authors:** Silke Gastine, Juanita Pang, Florencia A.T. Boshier, Simon J. Carter, Dagan O. Lonsdale, Mario Cortina-Borja, Ivan F.N. Hung, Judy Breuer, Frank Kloprogge, Joseph F. Standing

## Abstract

SARS-CoV-2 viral loads change rapidly following symptom onset so to assess antivirals it is important to understand the natural history and patient factors influencing this. We undertook an individual patient-level meta-analysis of SARS-CoV-2 viral dynamics in humans to describe viral dynamics and estimate the effects of antivirals used to-date. This systematic review identified case reports, case series and clinical trial data from publications between 1/1/2020 and 31/5/2020 following PRISMA guidelines. A multivariable Cox proportional hazards regression model (Cox-PH) of time to viral clearance was fitted to respiratory and stool samples. A simplified four parameter nonlinear mixed-effects (NLME) model was fitted to viral load trajectories in all sampling sites and covariate modelling of respiratory viral dynamics was performed to quantify time dependent drug effects. Patient-level data from 645 individuals (age 1 month-100 years) with 6316 viral loads were extracted. Model-based simulations of viral load trajectories in samples from the upper and lower respiratory tract, stool, blood, urine, ocular secretions and breast milk were generated. Cox-PH modelling showed longer time to viral clearance in older patients, males and those with more severe disease. Remdesivir was associated with faster viral clearance (adjusted hazard ratio (AHR) = 9.19, *p*<0.001), as well as interferon, particularly when combined with ribavirin (AHR = 2.2, *p=*0.015; AHR = 6.04, *p =* 0.006). Combination therapy should be further investigated. A viral dynamic dataset and NLME model for designing and analysing antiviral trials has been established.

## INTRODUCTION

Finding antivirals that target severe acute respiratory syndrome coronavirus 2 (SARS-CoV-2) will be crucial in managing the ongoing pandemic. In addition to the development of novel agents, substantial efforts are underway to establish whether currently available agents may be re-purposed^1^. A key biomarker for clinical antiviral activity is viral load in bodily fluids and assessing a drug’s or drug combination’s ability to reduce viral load is an important first step in identifying therapies that influence clinical outcome.

To correctly assess antiviral activity, it is first necessary to understand viral load natural history. As a rapidly progressing, primarily respiratory viral infection, SARS-CoV-2 elimination from the body seems to be mainly driven by a combination of innate immune response and exhaustion of target cells available for infection^2^. Observational cohort studies published to date have shown that the rate of viral load decline seems slower in older patients, those with more severe disease and those with comorbidities such as diabetes mellitus and immunosuppression^3, 4, 5, 6^. Interpreting these observational studies requires caution because patients have often received antiviral therapies. Due to the time point of initial infection being unknown, assessing viral load in response to treatment must account for time since symptom onset^7^.

Since February 2020 case reports and case series of patient-level viral dynamics have been published, some of which report dosing of antiviral drugs^8^. Clinical trials of antivirals and their association with viral load are also beginning to read out^9^. Meanwhile large pragmatic trials of repurposed monotherapy antivirals have yet to find a clearly effective agent^10^. At this crucial juncture, it is vital to develop a pharmacodynamic modelling framework that can be used to describe the natural history of SARS-CoV-2 viral dynamics, make initial estimates on antiviral efficacy of agents used to-date, and to design and evaluate Phase II trials using viral load as a biomarker.

This systematic review therefore aimed to search for case reports, case series and clinical trials reporting serial individual patient-level SARS-CoV-2 viral load measurements in humans from any sampling site upon which an individual patient-level meta-analysis was then performed. A nonlinear mixed effects (NLME) viral dynamic model was fitted to describe the viral trajectories in each sampling site and to give a quantitative measure of viral dynamics. In data of sufficient quality, the parameters of multivariable Cox proportional hazards regression models of time to viral clearance, and NLME models of antiviral efficacy were estimated.

## METHODS

### Protocol and registration

The protocol for this systematic review and individual patient meta-analysis, which follows the PRISMA Individual Patient Data systematic reviews guideline^11^, was first published on 27/5/2020 at: https://github.com/ucl-pharmacometrics/SARS-CoV-2-viral-dynamic-meta-analysis. The final dataset and statistical analysis code are also published here. The review was registered with PROSPERO (CRD42020189000).

### Eligibility criteria

This study aimed to identify serial viral loads with time in human subjects infected with SARS-CoV-2 in order to describe and model viral load trajectory. The inclusion criteria were therefore papers containing individual subject-level reports of viral load with time, either since symptom onset or time since start of monitoring for asymptomatic subjects, and sampling site. Authors of manuscripts describing summary statistics of viral load with time were contacted requesting participant level data. Viral load was defined as either a value in copies/mL or a cycle threshold (Ct) value of an uncalibrated polymerase chain reaction (PCR) assay.

### Overall search strategy

Since SARS-CoV-2 was notified to the WHO on 31/12/2019, we did not expect to find relevant papers published prior to this date. Hence, PubMed, EMBASE, medRxiv, and bioRxiv were searched with a date range of 1/1/2020 to 31/5/2020. The following search terms were used for PubMed and EMBASE: (SARS-CoV-2 OR COVID OR coronavirus OR 2019-nCoV) AND (viral load OR cycle threshold OR rtPCR OR real-time PCR OR viral kinetics OR viral dynamics OR shedding OR detection OR clinical trial). Due to character limits in the search engine, the following search terms were used for medRxiv and bioRxiv: (SARS-CoV-2 OR COVID-19 OR coronavirus) AND (viral load OR cycle threshold OR PCR OR viral dynamics OR clinical trial).

After removing duplicates, two reviewers independently identified papers for full text screening, with any discrepancies resolved by a third reviewer.

### Data extraction

Viral loads were reported as either numerical values in tables, figures, or in viral load *versus* time plots. Where possible, numerical values were copy-pasted directly into a comma separated value (csv) format from the source, whereas tabulated numerical values contained in pdf images were extracted using https://extracttable.com/. Viral loads reported in plots were extracted using Web Plot Digitizer^12^.

Each viral load was paired with a time since symptom onset or in asymptomatic subjects, the time since viral monitoring started. Furthermore, sampling site and, if viral load not reported in copies/mL, the PCR assay including the primers used, were extracted along with limit of quantification and limit of detection, if available. The following patient-level covariates were extracted if available:

- Presence of fever >37.5 °C at any time (non-time varying covariate)
- age, where possible individual age but otherwise the study’s reported central measure (e.g. mean, median)
- sex or the male/female ratio was extracted if patient-level data not reported
- need for and days of intensive care treatment
- need for and days of mechanical ventilation
- whether patient died and time to death from symptom onset.

In addition, a standardised disease score was constructed for each patient as follows:

0- asymptomatic
1- mild disease (fever, cough or other mild symptoms reported)
2- moderate disease (in addition to mild criterion: need for supplemental oxygen /non-invasive ventilation)
3- severe disease (requirement for mechanical ventilation)

All data were stored on a shared github repository, and standardised R-scripts took data from each paper to merge into a single master dataset. A quality control (QC) check on viral load values and all covariates was performed for each paper by an independent reviewer.

### Data quality assessment

#### Viral load quality score

Two quality assessments were applied to each dataset. Firstly, the quality of viral load reporting was rated on a 1-3 scale. The highest quality 1 was assigned to studies reporting viral load in copies/mL or reporting a calibration curve allowing for direct conversion of Ct values to viral load. Quality 2 was assigned if viral load was reported in PCR Ct and primers used in the assay were reported, but calibration data was missing. In this case a published calibration curve for that primer from another source was used to convert to viral load in copies/mL^13, 14^. Where more than one calibration curve was available for the same primer the mean slope and intercept was used. The lowest (quality score 3) was assigned when viral load was reported in PCR Ct but no further information was available on the PCR assay. In this instance a conversion to copies/mL was made using the mean slope and intercepts from all calibration curves.

#### Drug quality score

The second quality assessment on a 3-point scale related to reporting of the antiviral drug therapy administered: which drug(s) and upon which days did patients receive the drug(s). The highest quality 1 was assigned when it was reported which days each patient received each drug, or these data were provided by corresponding authors. If it was reported that no antiviral was administered this was also assigned quality 1. Quality 2 was assigned when antiviral drug treatment was reported, but ascertaining which days the patient had received the drugs was not possible. The lowest category, quality 3, was assigned when it was not possible to determine whether or not antivirals had been administered.

### Statistical analysis

#### Primary analysis of time to viral clearance using Cox proportional hazards modelling

The primary analysis was conducted on observed time to viral clearance, which was analysed fitting Cox proportional hazards regression models with adjusted hazard ratios estimated for each covariate. We verified the assumptions of proportional hazards using the Therneau-Grambsch test.^15^ The data used for this analysis were limited to respiratory and stool sampling sites only, as virus was found to be mostly undetectable at other sites. Furthermore, only data from patients with known antiviral history (drug quality 1 and 2) were used. To assess the possible risk of bias in different drug and viral load qualities, the analysis was repeated on two further subsets: Firstly, with only drug quality 1 and respiratory samples, and secondly on assay quality 1 data only.

Time to viral load dropping below the limit of detection was modelled with Cox proportional hazards regression in R (version 3.6.3)^16^. Where a single patient contributed samples from multiple sampling sites (e.g. upper respiratory and stool), the time to the last site testing negative was used. Multivariable models for covariate effects on time to viral clearance were fitted, with additional interaction terms for drug therapies included, where multiple antiviral agents were given simultaneously. In studies reporting sex as a proportion of males, 10 000 datasets were simulated using the reported fraction of males to randomly assign individuals to being male from the binomial distribution. The Cox proportional hazards regression model was then fitted to each dataset and parameter estimates compared with the model, where individual sex was assigned by rounding the fraction of males. Model parameter estimates were visualised using forest plots.

#### Secondary analysis antiviral pharmacology model

The secondary analysis was to use a NLME model to quantify the increase in viral elimination rate with antiviral therapy. This analysis used data only from respiratory samples and rated drug quality 1.

#### Nonlinear mixed-effects (NLME) viral dynamic model

Firstly, a descriptive analysis of all data was undertaken. A NLME viral dynamics model was fitted to the individual patient-level viral load *versus* time data. The structural model was based on the general target cell limited model, which has previously been used to describe respiratory viral infections^7, 17^. This model consists of three ordinary differential equations relating to changes in uninfected target cells (*T*), infected target cells (*I*) and free virus (*V*) over time (*t*), as follows:

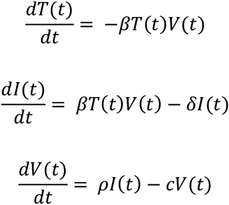

where *β* is the rate at which target cells become infected in the presence of virus, *δ* is the death rate of infected cells, *ρ* is the rate of viral production from infected cells and *c* is the rate of clearance of free virus. This model is structurally unidentifiable, as tested through the *IdentifiabiltyAnalysis* package in Wolfram Mathematica 12.1 (Wolfram Research, Illinois, USA) ^18^, unless the initial condition for *T, β*, or *ρ* are known. Furthermore, the elimination rate of free virus (*c*) is likely to be much faster than the death rate of infected cells (*δ*). Hence, by assuming a quasi-steady-state between *I* and *V*, and normalising the total cell number by the number of infected cells when observations begin (*t* = 0), it is then possible to reduce the model to a structurally identifiable, two state ordinary differential equation model relating to the fraction (*f*) of infected cells with time and infected cells as a proxy for viral load as follows^19^:

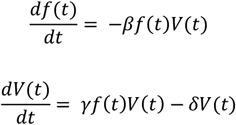

with *γ*, a new parameter equal to *ρβT*_*0*_*/c* and interpreted to be the maximum rate of viral replication. *δ* can now be interpreted as overall viral elimination rate. This population model was then fitted to viral load data with time using the following form:

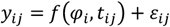

where *y*_*ij*_ was the viral load from subject *i* at time *t*_*ij*_, *f* is the nonlinear model defined above with parameters *φ*_*i*_, and *ε*_*ij*_ the residual between the model prediction and the observed data.

Four parameters were estimated: the initial viral load at symptom onset (*V*_*0*_), *β, δ* and *γ*. Interindividual variability was estimated for *V*_*0*_, *β* and *δ* with each assumed to follow a log-normal distribution. Viral loads were log transformed and the residual error was assumed to follow a normal distribution. Parameter estimation by maximum likelihood was undertaken using the stochastic approximation expectation maximization (SAEM) in NONMEM version 7.4^20^. Model evaluation was undertaken by analysis of normalised prediction distribution errors (NPDE) and visual predictive checks (VPC)^21^. Viral loads below the limit of detection (LOD) were included by integrating the density function from minus infinity to the limit of detection to yield a probability of the data being below the LOD (“M3 Method”)^22^.

In some participants, multiple samples were taken at the same time point (either different sampling site or the same sample assayed by more than one method). In this case a common residual error term was used to allow for modelling one-level nested random effects.

#### Descriptive analysis of viral shedding by sample site

The above model was fitted to data from each sampling site. The resulting parameters were then used to simulate the overall population viral load trajectories. For the respiratory sample sites viral area under the curve (AUC), peak viral load and half-life were derived from the model and plotted *versus* patient covariates.

#### Covariate analysis and antiviral drug effects modelling

The initial model used only data obtained in untreated patients. A covariate analysis was undertaken testing the influence of sampling site (nasal *versus* oral *versus* lower respiratory tract), sex, age and disease status on either *V*_*0*_, *β* or *δ*. Covariates were retained in the model based on the likelihood ratio test with a threshold level of significance of *p*<0.01, and if the same covariate addition to *V*_*0*_, *β* or *δ* all gave significant improvement to model fit then the model with the largest decrease in -2 log likelihood (NONMEM objective function value (OFV)) was chosen. For the final model viral area under the curve (AUC), peak viral load and half-life were derived and plotted versus patient covariates.

Using the final demographic model, data from patients undergoing antiviral treatment (antiviral drug quality 1) were added. A univariable analysis was performed, testing each drug’s ability to increase *δ*. Drugs showing significant improvement in model fit (p<0.01), according to the likelihood ratio test, were then included in the final multivariable model.

### Simulations based on the antiviral pharmacology model

Simulations were performed to explore the change in viral trajectories for different time points of therapy initiation: Day 1 after symptom onset, Day 3, Day 7 and Day 10. Interferon and ribavirin monotherapy along with the combination therapy interferon plus ribavirin were explored this way. A dummy population of 5100 subjects with ages uniformly distributed across 50 to 100 years, consisting of an equal ratio of males and females was created. Each regimen was simulated using the entire population, assuming sampling from the upper respiratory tract or nose for a time window of 14 days. Comparisons of the sample size required to detect a significant difference in the proportion of undetectable virus between antiviral and no treatment were made after 7 days of treatment with a 90% power and alpha level of *p*<0.05 for antivirals starting at Days 1, 3 and 7 post symptom onset.

## RESULTS

Results of the systematic search are given in Figure 1, and details of included papers in Table 1. Individual patient-level data were extracted from 45 articles reporting viral loads and/or PCR Ct values with time since symptom onset. Of these 32 papers either reported antiviral participant-level drug histories, or these were provided by the corresponding author. The full dataset contained 645 individuals contributing 6316 viral load samples. The majority of samples (*n*) were taken from the respiratory tract: nasopharyngeal (315 individuals, *n=*2208), oropharyngeal or saliva (381 individuals, *n=*2144) and lower respiratory tract (81 individuals, *n=*799). The other reported samples sites were stool/rectal swabs (99 individuals, *n=*655), blood/plasma (42 individuals, *n=*258), urine (31 individuals, *n=*112), ocular (16 individuals, *n=*50), breastmilk (4 individuals, *n=*90). Metrics of the full data set are given in Supplementary Table S1.

**Table 1:**
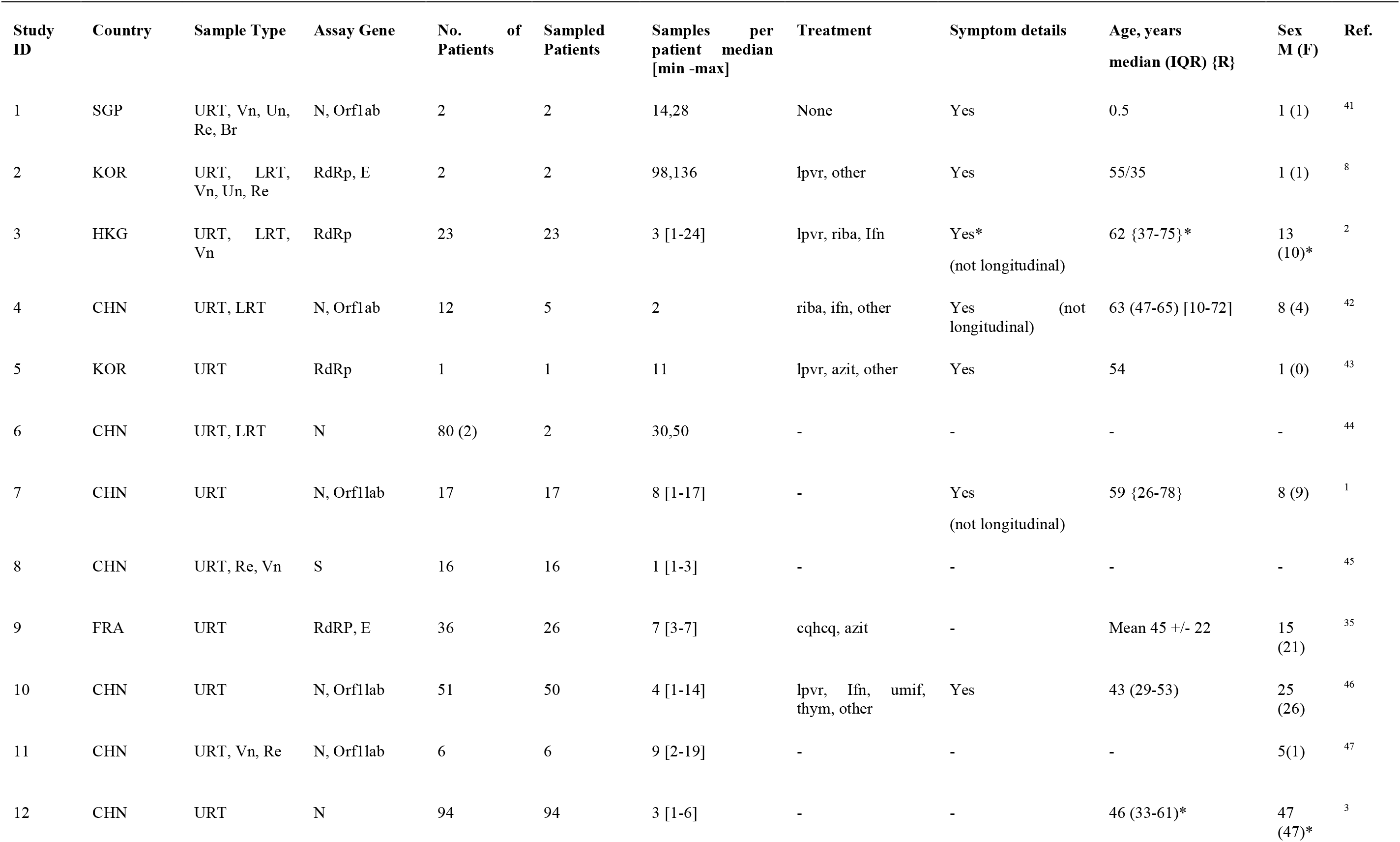
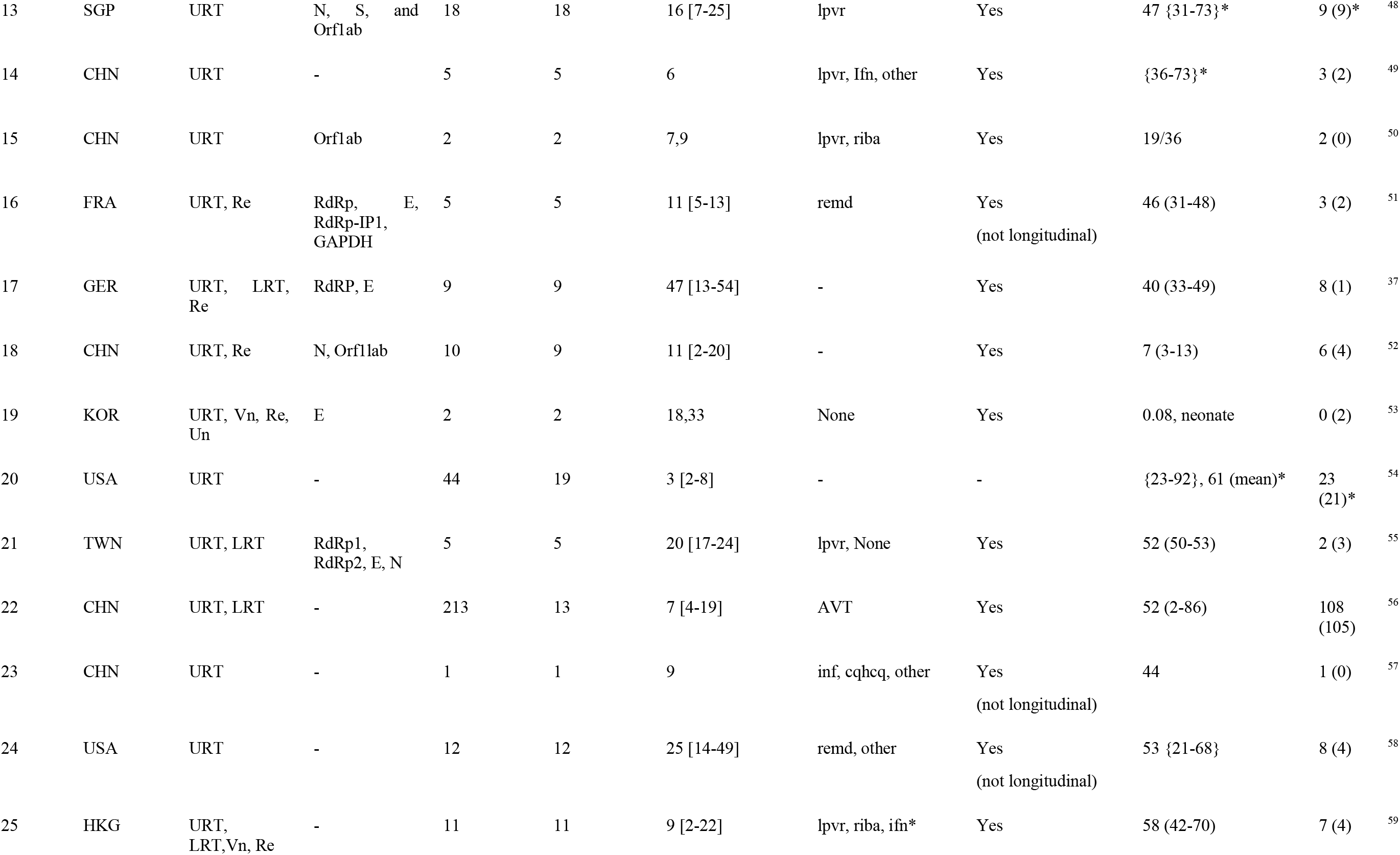
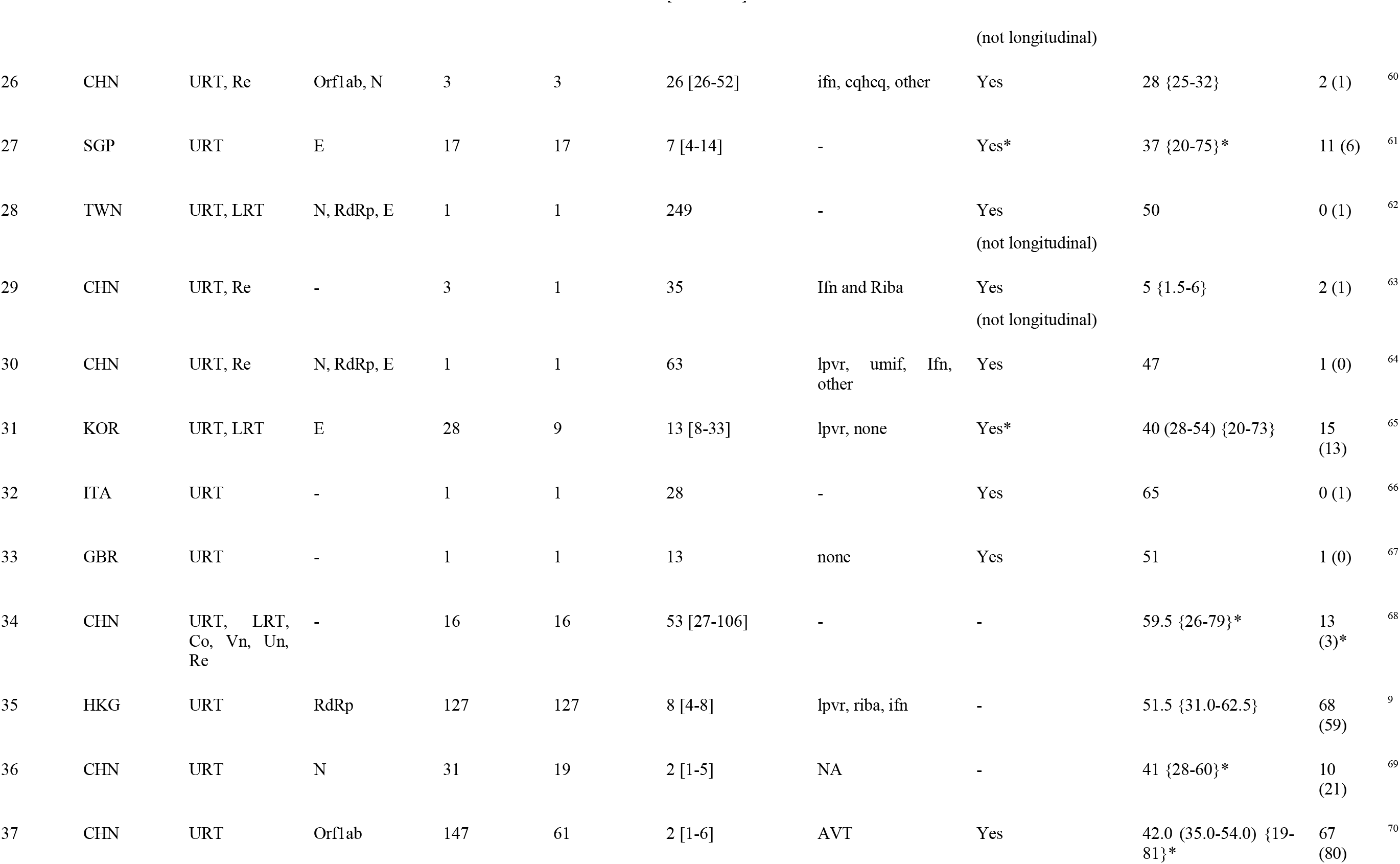
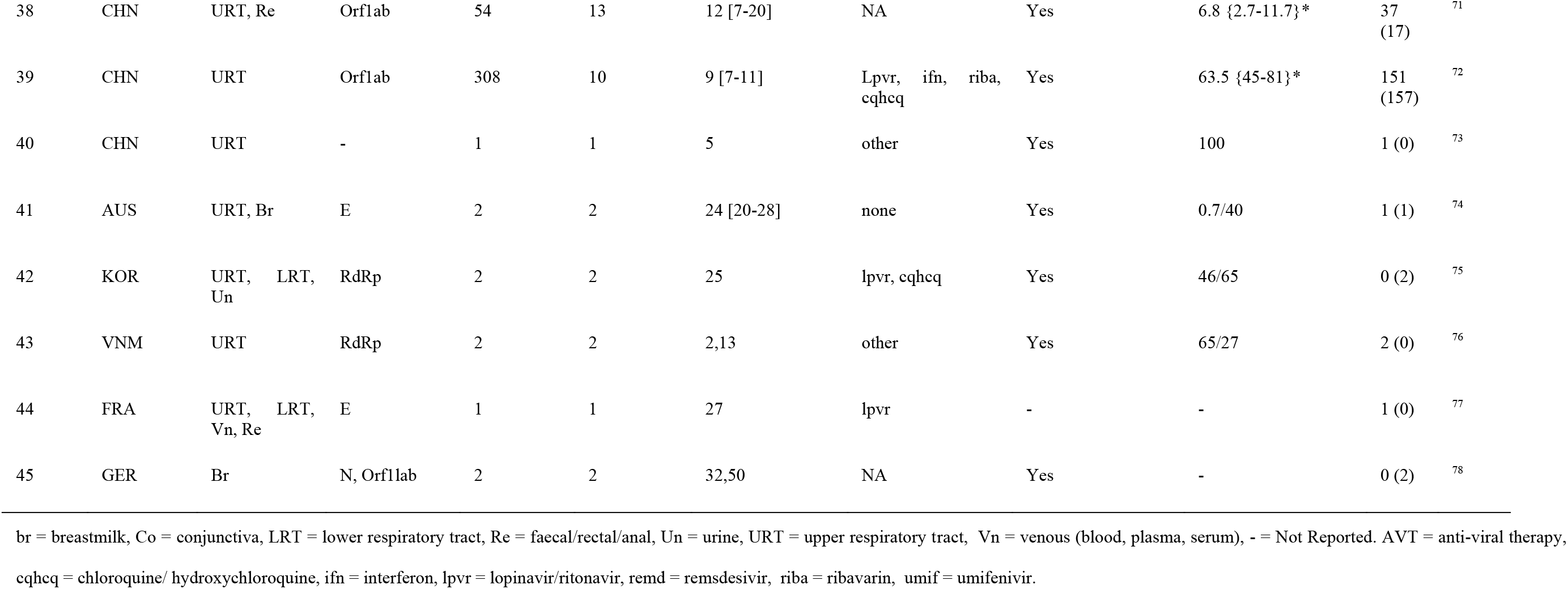
Individual papers included in the Meta-Analysis

**Figure 1:**
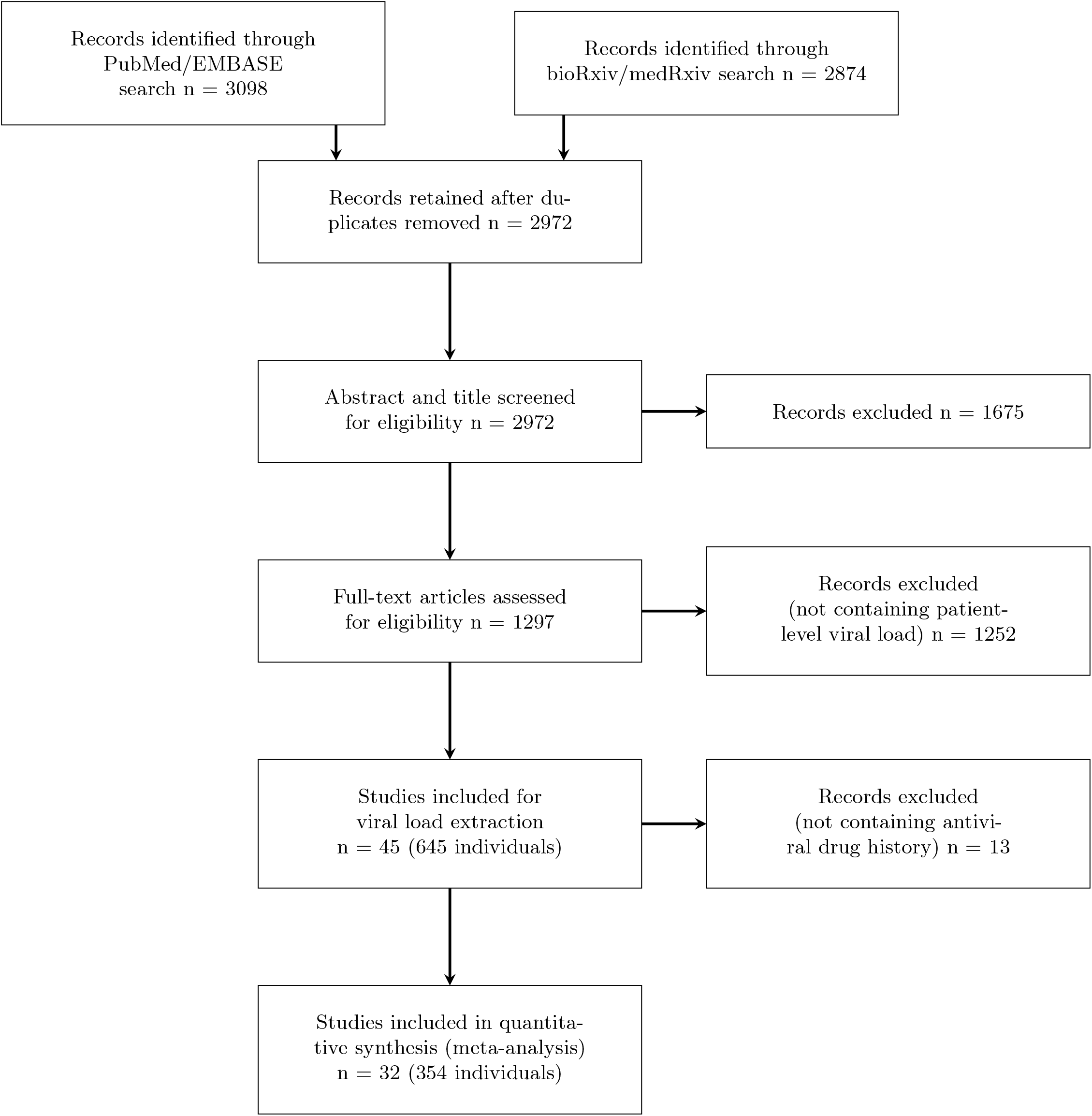
PRISMA diagram detailing the systematic search results.

Full details of the extracted patient-level covariates are given in Table 2. Recording of fever, days on ICU and days ventilated was largely unavailable. Therefore, no further analysis was performed on these variables. However, it was possible to categorise disease status in all drug quality 1 and 2 papers, either through reports in the manuscript or by contacting corresponding authors. Overall, most patients had mild disease 376 (66.8%), whereas 79 (14.0%) patients had moderate and 84 (14.9%) severe disease. In total 24 (4.3%) asymptomatic patients were reported. The distribution of recorded drug therapies, available for drug quality 1 data and respiratory site samples, is summarised in Supplementary Table S2. Sixty-seven of these patients did not receive antivirals.

**Table 2:** Overview extracted variables across different analyses, median [range] (%missing data records). n, number of individuals included.

| Descriptive (%missing) | All Data<br>*n*=645 | Cox-PH - Full data<br>set<br>*n*=354 | NLME/ reduced<br>Cox-PH<br>*n*=317 |
| --- | --- | --- | --- |
| Age [years] | 46 [0.1 – 100]<br>(31.3%) | 48 [0.1-100]<br>(0%) | 46 [0.1-100]<br>(0%) |
| Sex [male/female] | 217/189<br>(37%) | 215/139<br>(0%) | 182/135<br>(0%) |
| ICU admission [yes/no]* | 36/371<br>(36.9%) | 8/271<br>(21.2%) | 8/257<br>(16.4%) |
| Invasive ventilation [yes/no]* | 14/348<br>(43.9%) | 9/262<br>(23.4%) | 5/247<br>(20.5%) |
| Death [yes/no] | 1/455<br>(29.3%) | 1/330<br>(6.5%) | 1/293<br>(7.3%) |
| Disease severity* | (12.7%) | (0%) | (0%) |
| Asymptomatic | 24 | 19 | 16 |
| Mild | 376 | 258 | 239 |
| Moderate | 79 | 52 | 44 |
| Severe | 84 | 25 | 18 |

\*There is discord between the reported ICU and mechanical ventilation and disease severity score due to incomplete reporting in some papers. Disease severity was taken from individual reports of disease status in cases where ICU admission and invasive ventilation were not specifically mentioned, and only Disease severity was used in the analyses.

The NLME model fits to the overall data, stratified by sampling site, are provided in Supplementary Table S3 and Supplementary Figure S1. Simulations from the models for each sampling site showing the expected viral load trajectory along with the predicted proportion of samples, that would be below the limit of detection are given in Figure 2. For respiratory sites model-derived AUC, peak viral load and half-life is given in Supplementary Figure S2

**Figure 2:**
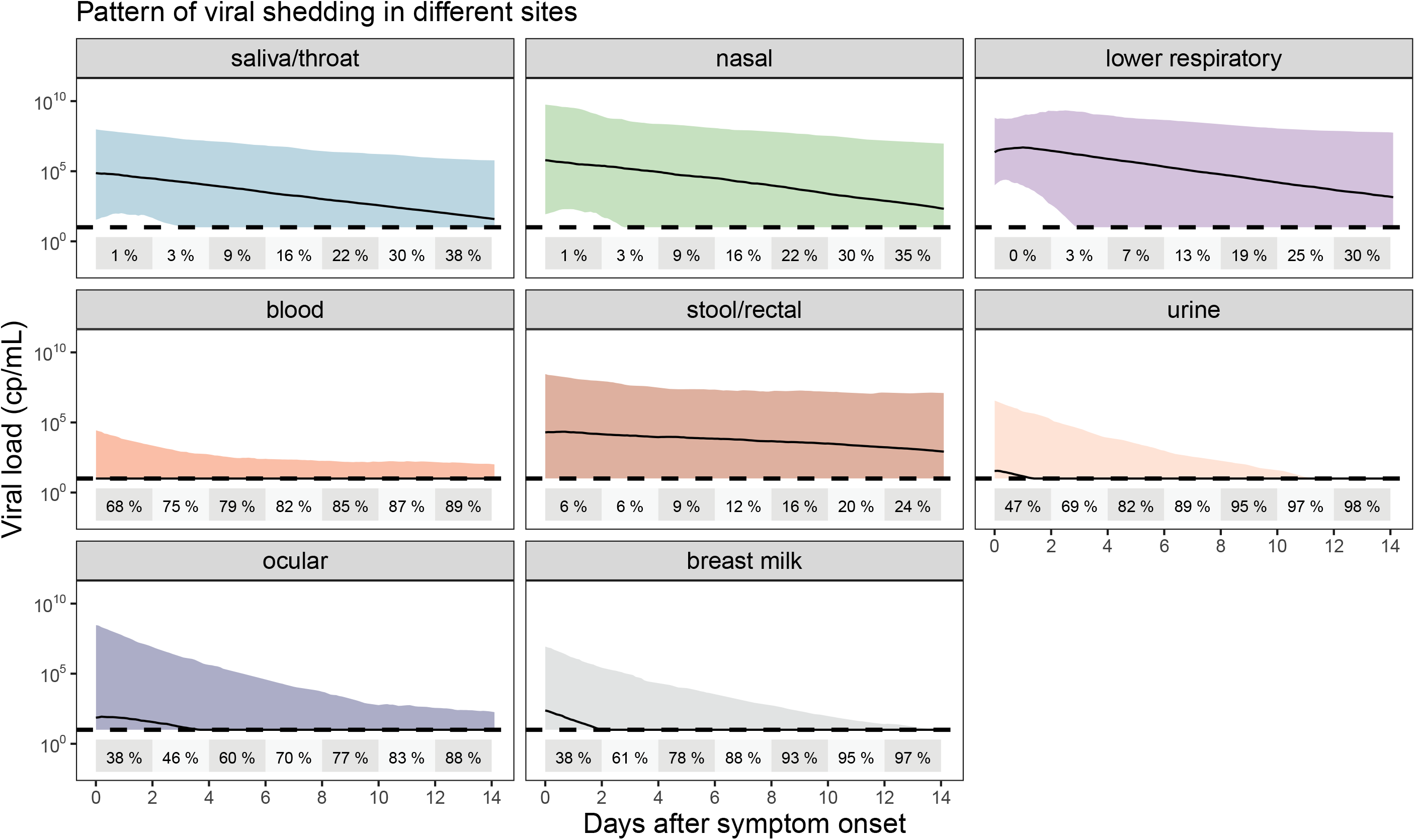
Model-predicted viral load trajectories at each sample site studied. Black lines are the median predictions, with shaded areas representing the 95% prediction interval. The percentage of samples that are predicted to be below a typical limit of detection (10 copies/mL) are given in 2-daily time bins on each plot.

Data on a total of 354 patients with respiratory and/or stool/rectal sampling and drug quality 1 or 2 were available. A forest plot of the parameter estimates from the Cox proportional hazards regression model is provided in Figure 3. Viral clearance was fastest from upper respiratory tract samples and slowest from stool. More sensitive assays (with lower detection limits) were associated with longer time to viral clearance and viral clearance was faster in females, younger patients and those who were asymptomatic.

**Figure 3:**
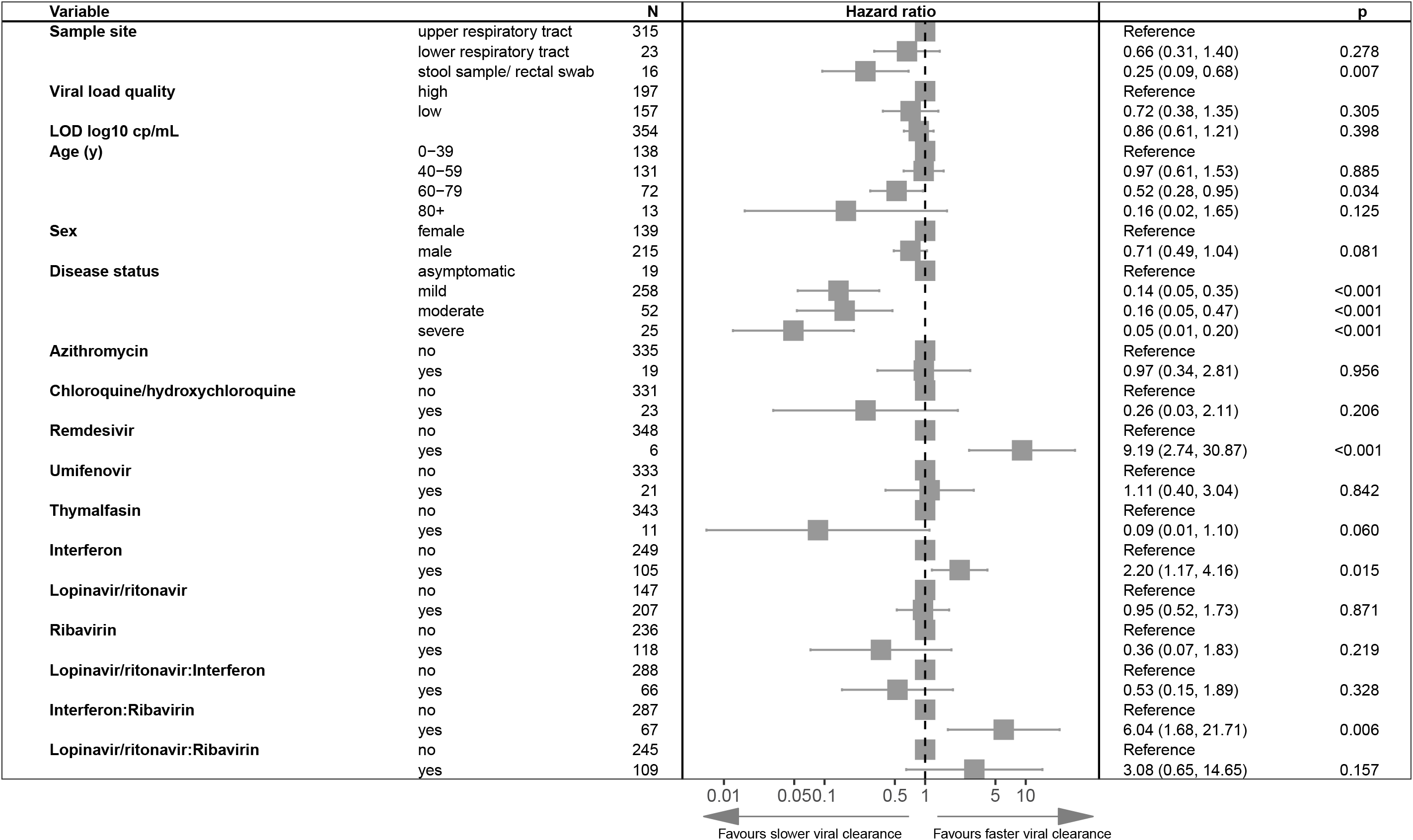
Multivariable Cox proportional hazard results on all drug quality 1 and drug quality 2 data from respiratory and stool/rectal sampling sites. Adjusted hazard ratios exceeding 1 indicate virus being more likely to become undetectable.

Regarding antiviral therapies, only remdesivir (adjusted hazard ratio (AHR) = 9.19, p<0.001) and interferons (AHR = 2.20, p =0.015) were independently associated with faster viral clearance. The effect of interferon alpha and beta (Supplementary Figure S3) was similar and hence these were combined. Lopinavir/ritonavir, ribavirin and interferons were most used and also most used in combination. Adding interaction terms for interferon plus lopinavir/ritonavir, interferon plus ribavirin and lopinavir/ritonavir plus ribavirin in the Cox proportional hazard regression analysis showed a trend towards synergy between interferons and ribavirin in the full dataset (AHR = 6.04, p=0.006 Figure 3), as well as in the additional analysis taking in quality assessments to account for potential bias: respiratory data limited to drug quality and in data limited to only viral load quality 1 data (Supplementary Figure S4 and Figure S5). Median sampling frequency in the main survival dataset was 1 day but there was a skewed distribution of sampling frequencies with the mean being 1.9 days and 4.8% of sampling frequencies being greater than 3 days. The main analysis was repeated excluding events with sampling frequencies over 3 days to check for potential bias caused by interval censoring, but the main effect sizes were similar (Figure S6).

Covariate relationships and drug effects were explored through NLME modelling with parameter estimates of the model given in Supplementary Table S4 along with visual predictive checks and NPDEs in Figure 4 and Supplementary Figure S7 and visualization of viral area under the curve, peak viral load and half-life derived from the final model in Supplementary Figure S8. Drug effects were estimated to increase *δ*. Drug regimens containing interferon (ΔOFV = -25.5, p<0.001), lopinavir/ritonavir (ΔOFV = 9.97, p=0.0016) and ribavirin (ΔOFV = -22.2, p<0.001) each improved model fit and so were taken forward to the final multivariable drug model. The estimated small lopinavir/ritonavir effect on delta, although showing significant model improvement in the likelihood ratio test, did not prove to be robustly detected in the bootstrap analysis, with the interval crossing the value consistent with no drug effect (Table S4). Implementing an additional synergy term, as detected in the Cox proportional hazard model, did not improve the NLME model.

**Figure 4:**
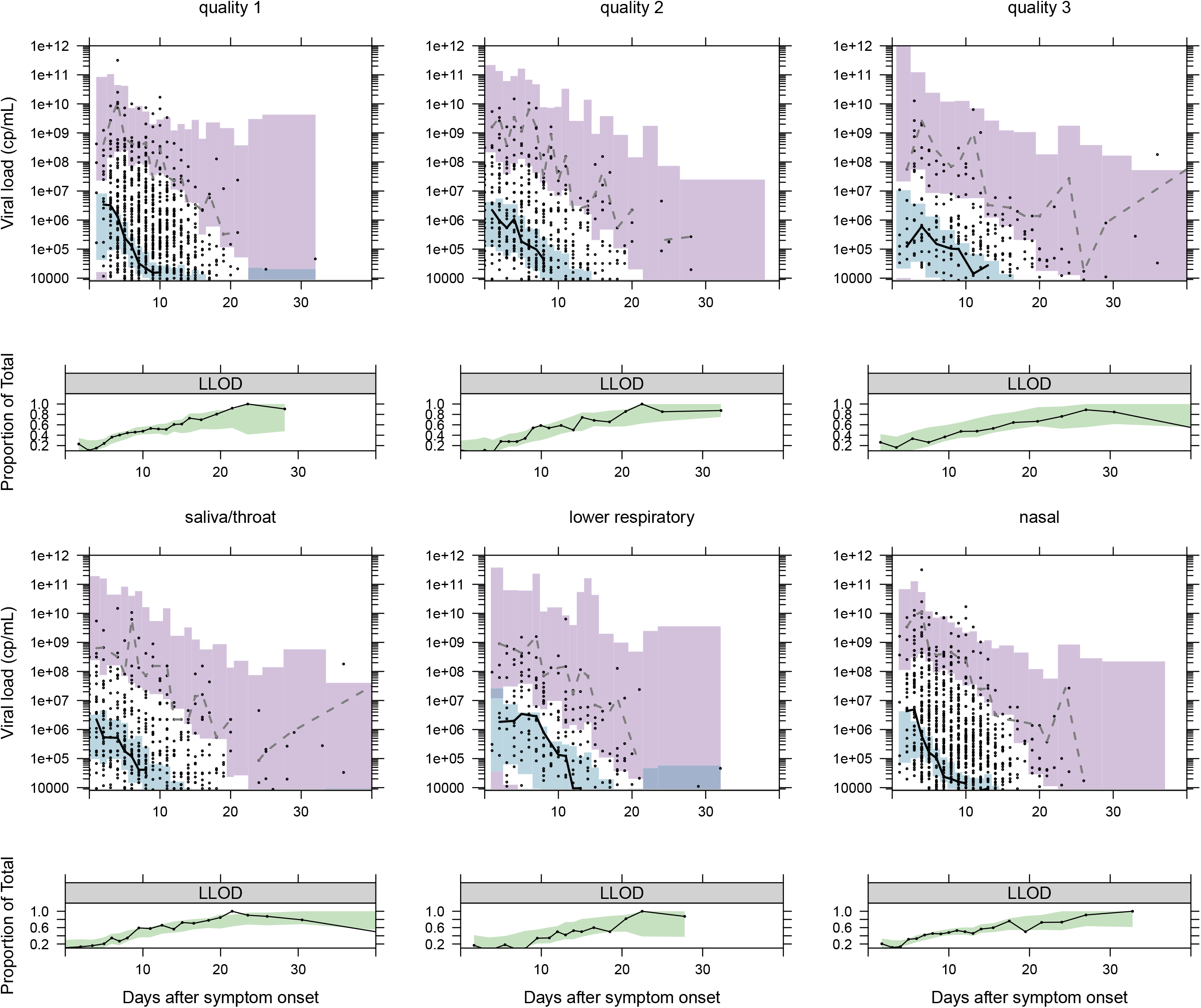
Visual predictive checks for the NLME model fitted to viral load data to each sampling site. For each site a plot of model simulations compared with observations is given for both the continuous data (upper) and the fraction of samples below the limit of detection (lower). Black circles are observed viral loads, purple shaded area is the 95% prediction interval of the simulated 2.5th and 97.5th percentile for comparison with the observed 2.5 and 97.5th percentile (dashed lines). The blue shaded area is the 95% prediction interval of the 50th percentile to compare with the continuous black line. In the lower plot the observed proportion of samples below the lower limit of detection (LLOD) are shown as a black line and compared with the 95% prediction interval of the model predicted proportion of samples below the LLOD (green shaded area).

The final model was then used to simulate expected viral trajectories from upper respiratory sampling sites for interferon, and ribavirin monotherapy as well as interferon plus ribavirin combination started at 1, 3, 7 and 10 days post symptom onset (Figure 5). The sample sizes for hypothetical Phase II trials to detect significant differences in viral load versus no treatment after 7 days of therapy are given in Supplementary Table S5.

**Figure 5:**
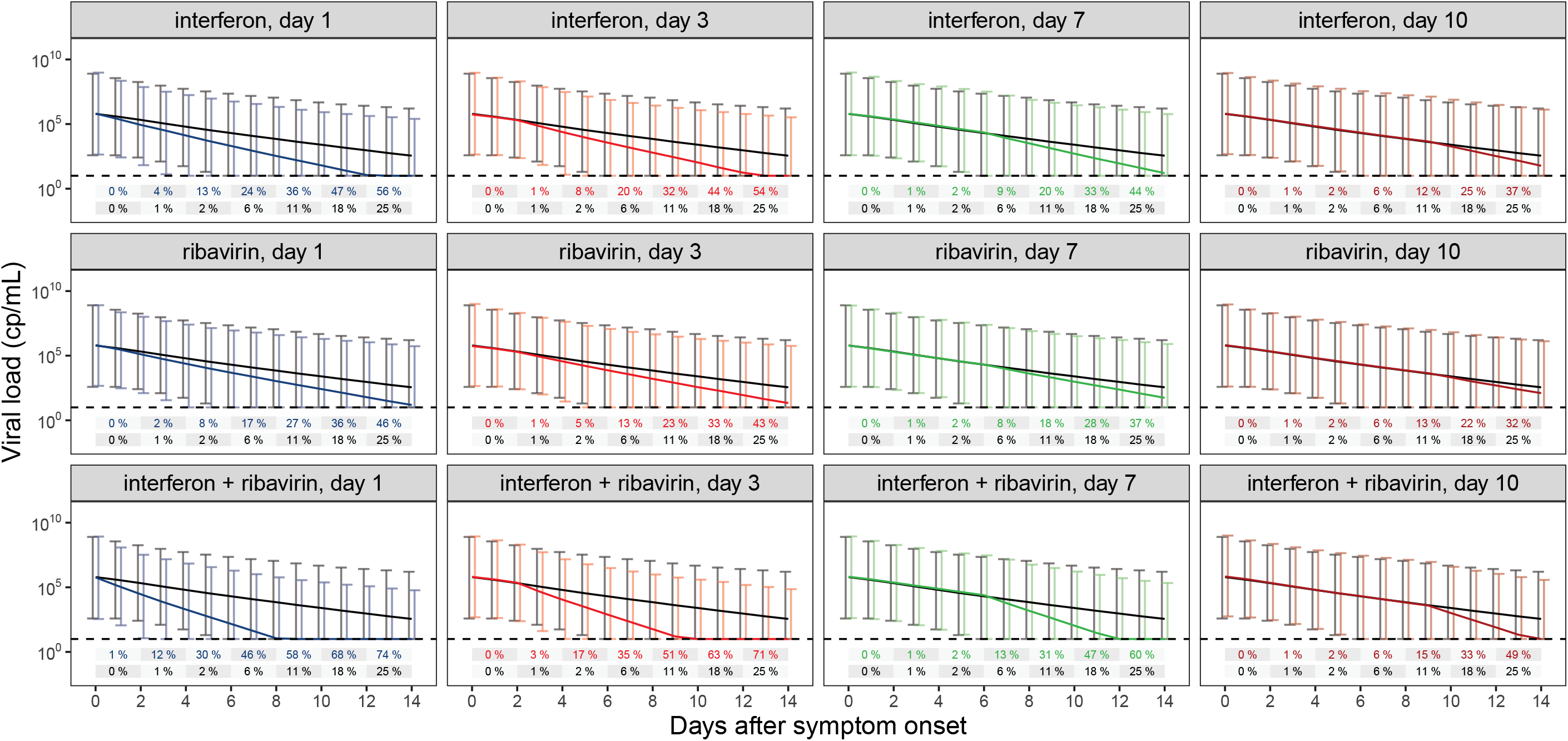
Simulated viral load trajectories. Simulations with a dummy population equally distributed between 50 and 100 years, and equal male/female ratio were performed for each scenario. Drugs were started at day 1 (blue), day 3 (orange), day 7 (green) or day 10 (red) post symptom onset. Mean black line and error bars represent simulations of the dummy population without drug treatment. Coloured mean lines and error bars represent the respective drug regimen. Percentage values represent expected proportion of samples below the limit of detection for no drug (black) versus drug therapy (coloured) at each time point.

## DISCUSSION

This systematic review and individual level meta-analysis has identified viral load trajectories from 645 individuals aged from the first month of life to 100 years. Data from all major sampling sites showed, that: following symptom onset in most patients, upper respiratory tract viral load has peaked and is declining, whereas in the lower respiratory tract viral load peaks 2-3 days after symptom onset; virus is detectable in stool for at least 2 weeks in 75% of individuals, and virus is detected in low levels in blood, urine, ocular secretions and breast milk (Figure 2). In addition to simulating the expected trajectory of viral load at each site, we were able to simulate the percentage of samples expected to be below a typical detection limit of 10 copies/mL (Figure 2). From this it can be seen, that from day 10 post symptom onset over a quarter of upper respiratory samples have undetectable viral load. This emphasises the importance of early antiviral therapy, and for Phase II trials using viral load as an endpoint to commence therapy in the first few days of symptom onset in order to reliably differentiate antiviral effects from natural viral decline (Figure 5, Supplementary Table S5).

Although we followed PRISMA guidelines on individual patient-level meta-analysis methods, registered our review with PROSPERO and prospectively published our analysis protocol prior to finalising our search, by including data from case reports, case series and clinical trials it could be argued that the heterogeneous inclusion criteria of these data may bias the treatment effects we estimated. We therefore repeated the primary analysis on subsets of the data based on sampling site, data quality and sampling frequency (Supplementary Figures S4-6) finding that the main effects were consistent. It should be noted that by far the largest drug quality 1 dataset was the clinical trial from Hung et al^9^ with 127 patients randomised to either lopinavir/ritonavir versus lopinavir/ritonavir plus ribavirin plus interferon β, and our second largest drug quality 1 group was those confirmed to have received no antiviral drugs (67 patients). In total our NLME dataset contained data on 83 patients receiving interferons, 187 patients receiving lopinavir/ritonavir, and 99 patients receiving ribavirin either alone or in combination (Table S2). Therefore, whilst consistency in results with various subgroup analyses indicate confounding related to heterogeneous data is unlikely to have biased our main findings, the heterogeneity in drug and drug combination studies meant modelling was required to tease out individual drug effects.

A heterogeneous range of antivirals, administered in different combinations, was observed in our data (Supplementary Table S2) meaning multivariable modelling of time to viral clearance was used to tease out individual drug effects. No antiviral activity was seen for chloroquine/hydroxychloroquine, azithromycin, lopinavir/ritonavir, umifenovir and thymalfasin. However, remdesivir and interferons were both independently associated with shorter time to viral clearance (Figures 3, S4 and S5). Remdesivir did not however significantly decrease *δ* in the NLME model, but this is likely due to the low number of included patients.

Our most interesting finding is the promising antiviral activity of interferons, possibly due to low endogenous interferon levels induced by SARS-CoV-2^23, 24^. Interferons (alpha and beta) have shown extensive *in vitro* activity against Severe acute respiratory syndrome-1 (SARS-CoV-1) and Middle East respiratory syndrome coronavirus (MERS-CoV)^25, 26^.However, this has not translated into clinical effectiveness in MERS-CoV^25^, although results from one trial are still pending^27^. Although recent data suggests interferon beta may be more potent than alpha against SARS-CoV-2 *in vitro*^28^, possibly due to higher selective indices for interferon-beta 1b, upon finding similar effects of interferon alpha and beta in our primary analysis (Figure S3), we decided to combine the interferon effect to better explore drug combinations. In the Cox proportional hazard analysis, consistent across data qualities and sampling site combinations, we found either a significant or trend towards significant synergistic activity of interferon plus ribavirin (Figures 3, 4, S4). An extensive body of literature exists to show both interferon alpha and beta are synergistic with ribavirin *in vitro* against both SARS-CoV-1 and MERS-CoV^25^. This synergy, however, was not confirmed when tested in the NLME analysis, indicating that the detected synergistic effect from the time to viral clearance analysis might be confounded. Correlations in the timing for start of drug treatment could be one confounder that is corrected for in the NLME approach. The time-dependant analysis from the NLME model suggests and additive effect for interferon and ribavirin, rather than a synergistic effect. Thus, combining interferons with a nucleoside analogue, possibly remdesivir or favipiravir as less toxic alternatives to ribavirin, is a potentially promising combination for viral load suppression. In our secondary analysis, we included interferon plus ribavirin in the NLME model and simulations show that virus should be suppressed 2-3 days faster compared to no treatment (Figure 5). However, it must be noted that recent evidence from the WHO SOLIDARITY trial shows that interferons were associated with a trend to increased mortality ^29^ whereas an unpublished press release reports inhaled interferon-β to be beneficial^30^. There is a clear need for a well-designed Phase II trial on interferons in early disease to confirm or refute the signal seen in our data.

Another main finding of our work was the limited antiviral effect of lopinavir/ritonavir, in addition to its lack of significant synergistic effect with either ribavirin or interferons. The protease inhibitor lopinavir had a modest but consistent *in vitro* activity against the major coronaviruses, including SARS-CoV-2, although activity is confined to concentrations at the upper end of the clinically achievable range^1^. Whilst lopinavir significantly improved model fit when increasing *δ*, the bootstrap lower boundary crossed the threshold of no drug effect (Table S4), and our simulations suggest monotherapy studies would require well over 500 participants per arm just to show antiviral activity. As recent Phase III trials have now conclusively shown, lopinavir/ritonavir is ineffective in monotherapy^29, 31^. It remains to be seen whether lopinavir/ritonavir may be useful in combinations, however. In SARS-CoV-1 lopinavir/ritonavir plus ribavirin was found to be synergistic *in vitro* and when initiated immediately upon diagnosis led to a significant decrease in mortality compared to historical controls^32, 33^. Early post-exposure prophylaxis against Middle East Respiratory Syndrome (MERS-CoV) in healthcare workers showed that lopinavir/ritonavir plus ribavirin reduced the incidence of infection from 28% to 0%^34^. The lopinavir/ritonavir plus ribavirin combination has therefore been the basis for many clinical trials and treatment protocols, but our findings suggest that it may not be as useful in SARS-CoV-2 (Figure 3).

The antiviral effects of remdesivir *in vitro* are well established and despite only being able to extract individual patient-level data on six patients, it produced a significantly faster viral clearance in the primary analysis (Figure 3). Despite in some cases showing promising *in vitro* activity, we did not find significant antiviral effects of azithromycin, chloroquine/hydroxychloroquine, thymalfasin or umifenovir. In the case of hydroxychloroquine and azithromycin the raw viral load data from the heavily criticised study by Gautret et al^35^ was included, but contrary to the original analysis we found no clinical antiviral activity of either drug and, in the case of hydroxychloroquine, a trend towards slower viral clearance. The reason for this difference in interpretation appears to stem from using time since symptom onset as opposed to time since starting drug and with untreated patients being monitored from an earlier day-post symptom onset. This example highlights the necessity of accounting for the time course of the infection when analysing viral loads.

In our secondary NLME analysis the simplified target cell limited model provided a good fit to data from each sampling site. In many cases this approximated a mono-exponential decay, but in others, particularly in lower respiratory tract, there was a pronounced peak in the first days following symptom onset. The model was stable with high inter-individual variability on V_0_ and β, reflecting the fact that relative changes in these parameters lead to the initial part of the curve either rising then falling (in situations when V_0_ ≈ -β) or approximately monoexponentially declining (when V(0) >> -β). In addition, we found the model to be less sensitive to changes in γ, meaning it can take a wide range of values with little influence on model fit, hence we did not estimate an inter-individual variability term on it. Increasing age was associated with significantly slower *δ*, and there was a small effect of male sex also being associated with slower *δ* (Table S4). The age effect translates to a 5-year-old having a viral decay terminal half-life of 1.0 day, a 47-year-old (median age in our population) 1.18 days and a 90-year-old 1.24 days. Hence a child has an almost 15% faster viral clearance than a middle-aged adult, and almost 20% faster than an elderly person.

In contrast to authors who have estimated parameters for more mechanistic models^36^, we estimated all drug effects to increase *δ*, which implies a mode of action relating to inhibition of viral replication or stimulation of viral clearance mechanisms. Whilst for most of the drugs studied this may be reasonable, entry inhibitors may be more appropriately described by inhibition of γ, which may not be statistically identifiable with the data possible to collect in the clinical setting. Despite this potential limitation, we found similar agents (combinations including interferons and ribavirin) to those identified in the primary analysis of time-to viral clearance.

The major limitation of our work is the lack of clinical trial data and lack of data on potentially important re-purposing agents such as favipiravir and nitazoxanide and that only one of the authors of a major clinical trial agreed to share their data^9^. Through applying quality assessment criteria on drug history and assay reporting, pre-specifying our analysis in our protocol and PROSPERO registration before undertaking Cox proportional hazards and NLME modelling we aimed to reduce possible bias in the heterogenous data available. Whilst we were able to extract a limited common demographics set, particularly in the high-quality data subset (age, sex, disease severity, antiviral drug histories), our data may be limited by other non-antiviral medications that were not fully reported in the included papers. Furthermore, as many of our included papers were on patients with mild or no symptoms and only contained data on one patient reported to have died, we were unable to study associations of viral load and mortality. Viral load measured by PCR is not necessarily infectious virus, and recently it has been shown that only in samples above 10^7^ copies/mL can SARS-CoV-2 be cultured^29^. Therefore, our data should preferably be used to study viral trajectories in relation to antiviral therapy rather than to infer probability of transmission.

The detection of viable virus might be overcome through whole genome sequencing and the detection of subgenomic RNA. This has however only been conducted in a single study, included in our review. Woelfel et al.^37^ showed through E gene subgenomic RNA quantification and relating it to the entire virus genome RNA, that presence of subgenomic RNA fragments can be a hint for active viral replication and thus active infection. More recent studies by Alexandersen et al^38^ and van Kampen et al^39^ however detected subgenomic RNA up to 22 days after onset of symptoms. It is postulated this was related to subgenomic RNA being rather stable and associated with cellular membranes and thus detection of subgenomic RNAs in clinical samples does not necessarily indicate viral activity. Future controlled studies of subgenomic RNA levels in patients on and off antiviral therapies are urgently required to better understand this potential biomarker of drug effect.

In conclusion, this individual patient level meta-analysis has yielded useful insights into SARS-CoV-2 viral dynamics. A model-based description of viral trajectories in different sampling sites has been elucidated, and we have found covariates such as increasing age, disease severity and male sex to be associated with slower viral clearance. Our review firmly establishes a role for early viral suppression in the management of SARS-CoV-2 and an important signal as to the possible benefits of interferons as a component of antiviral therapy has been found. It has been shown that viral dynamic models such as ours can increase the power to detect drug effects due to their utilisation of serial measures^40^ and our model should be useful to others in both the design and analysis of future Phase II trials, hence the model code and raw data from this analysis is made available.

## Data Availability

The final dataset is available at github

https://github.com/ucl-pharmacometrics/SARS-CoV-2-viral-dynamic-meta-analysis

## DATA AVAILABILITY

The final dataset is available at: https://github.com/ucl-pharmacometrics/SARS-CoV-2-viral-dynamic-meta-analysis

## CODE AVAILABILITY

Model code is available at: https://github.com/ucl-pharmacometrics/SARS-CoV-2-viral-dynamic-meta-analysis

## SUPPLEMENTAL FILES

1. Supplemental Material

