## Supplemental materials for "Systematic review and patient-level meta-analysis of SARS-CoV-2 viral dynamics to model response to antiviral therapies"

### PRISMA 2009 Checklist

| Section/topic | # | Checklist item | Reported on page # |
|---|---|---|---|
| **TITLE** | | | |
| Title | 1 | Identify the report as a systematic review, meta-analysis, or both. | 1 |
| **ABSTRACT** | | | |
| Structured summary | 2 | Provide a structured summary including, as applicable: background; objectives; data sources; study eligibility criteria, participants, and interventions; study appraisal and synthesis methods; results; limitations; conclusions and implications of key findings; systematic review registration number. | 2 |
| **INTRODUCTION** | | | |
| Rationale | 3 | Describe the rationale for the review in the context of what is already known. | 3 |
| Objectives | 4 | Provide an explicit statement of questions being addressed with reference to participants, interventions, comparisons, outcomes, and study design (PICOS). | 3 |
| **METHODS** | | | |
| Protocol and registration | 5 | Indicate if a review protocol exists, if and where it can be accessed (e.g., Web address), and, if available, provide registration information including registration number. | 5 |
| Eligibility criteria | 6 | Specify study characteristics (e.g., PICOS, length of follow-up) and report characteristics (e.g., years considered, language, publication status) used as criteria for eligibility, giving rationale. | 5 |
| Information sources | 7 | Describe all information sources (e.g., databases with dates of coverage, contact with study authors to identify additional studies) in the search and date last searched. | 5 |
| Search | 8 | Present full electronic search strategy for at least one database, including any limits used, such that it could be repeated. | 5 |
| Study selection | 9 | State the process for selecting studies (i.e., screening, eligibility, included in systematic review, and, if applicable, included in the meta-analysis). | 5 |
| Data collection process | 10 | Describe method of data extraction from reports (e.g., piloted forms, independently, in duplicate) and any processes for obtaining and confirming data from investigators. | 6 |
| Data items | 11 | List and define all variables for which data were sought (e.g., PICOS, funding sources) and any assumptions and simplifications made. | 6 |
| Risk of bias in individual studies | 12 | Describe methods used for assessing risk of bias of individual studies (including specification of whether this was done at the study or outcome level), and how this information is to be used in any data synthesis. | 7 |
| Summary measures | 13 | State the principal summary measures (e.g., risk ratio, difference in means). | 7-8 |
| Synthesis of results | 14 | Describe the methods of handling data and combining results of studies, if done, including measures of consistency (e.g., $I^2$) for each meta-analysis. | 7 |

### PRISMA 2009 Checklist

| Section/topic | # | Checklist item | Reported on page # |
|---|---|---|---|
| Risk of bias across studies | 15 | Specify any assessment of risk of bias that may affect the cumulative evidence (e.g., publication bias, selective reporting within studies). | 7 |
| Additional analyses | 16 | Describe methods of additional analyses (e.g., sensitivity or subgroup analyses, meta-regression), if done, indicating which were pre-specified. | 8-10 |
| **RESULTS** | | | |
| Study selection | 17 | Give numbers of studies screened, assessed for eligibility, and included in the review, with reasons for exclusions at each stage, ideally with a flow diagram. | 11 |
| Study characteristics | 18 | For each study, present characteristics for which data were extracted (e.g., study size, PICOS, follow-up period) and provide the citations. | 11, table1 |
| Risk of bias within studies | 19 | Present data on risk of bias of each study and, if available, any outcome level assessment (see item 12). | 11 |
| Results of individual studies | 20 | For all outcomes considered (benefits or harms), present, for each study: (a) simple summary data for each intervention group (b) effect estimates and confidence intervals, ideally with a forest plot. | 11, figure3 |
| Synthesis of results | 21 | Present results of each meta-analysis done, including confidence intervals and measures of consistency. | 12 |
| Risk of bias across studies | 22 | Present results of any assessment of risk of bias across studies (see Item 15). | 12 |
| Additional analysis | 23 | Give results of additional analyses, if done (e.g., sensitivity or subgroup analyses, meta-regression [see Item 16]). | 12 |
| **DISCUSSION** | | | |
| Summary of evidence | 24 | Summarize the main findings including the strength of evidence for each main outcome; consider their relevance to key groups (e.g., healthcare providers, users, and policy makers). | 13-15 |
| Limitations | 25 | Discuss limitations at study and outcome level (e.g., risk of bias), and at review-level (e.g., incomplete retrieval of identified research, reporting bias). | 16 |
| Conclusions | 26 | Provide a general interpretation of the results in the context of other evidence, and implications for future research. | 16 |
| **FUNDING** | | | |
| Funding | 27 | Describe sources of funding for the systematic review and other support (e.g., supply of data); role of funders for the systematic review. | 19 |

*From:* Moher D, Liberati A, Tetzlaff J, Altman DG, The PRISMA Group (2009). Preferred Reporting Items for Systematic Reviews and Meta-Analyses: The PRISMA Statement. PLoS Med 6(7): e1000097. doi:10.1371/journal.pmed1000097

For more information, visit: **www.prisma-statement.org**.

### Supplementary Material for “Systematic review and patient-level meta-analysis of SARS-CoV-2 viral dynamics to model response to antiviral therapies”

#### Supplementary Tables

**Table S1** Extracted viral load data metrics

| Sample site | *n* samples (%<LOD) | Viral load metrics [log10 copies/mL] | |
|---|---|---|---|
| | | Median (IQR) | [min – max] |
| **Blood** | 268 (79.9) | 3.96 (1.65) | 2.03 – 7.11 |
| **Saliva/Throat** | 2144 (37.1) | 4.62 (2.23) | 1.02 – 10.78 |
| **Lower respiratory** | 799 (28.9) | 5.47 (2.71) | 1.08 – 10.82 |
| **Stool/rectal** | 655 (35.4) | 5.05 (2.25) | -0.25 -11.38 |
| **Urine** | 138 (88.4) | 4.5 (1.22) | 3.76 – 6.35 |
| **Breast milk** | 90 (45.6) | 3.95 (2.63) | 1.96 – 5.31 |
| **Ocular secretion** | 50 (84.0) | 5.6 (1.99) | 2.85 -8.74 |
| **Nasal** | 2208 (20.8) | 5.00 (2.57) | 1.4 – 11.5 |

**Table S2** Summary of observed drug therapies – mono and combination therapies.

| **Drugs** | | **IDs** | **Samples** |
|---|---|---|---|
| | No drug | 67 | 862 |
| **Monotherapies** | | | |
| | Azithromycin | 1 | 1 |
| | Chloroquine/ hydroxychloroquine | 15 | 96 |
| | Interferon | 14 | 41 |
| | Lopinavir/ritonavir | 84 | 598 |
| | Remdesivir | 6 | 31 |
| | Ribavirin | 2 | 6 |
| | Umifenovir | 16 | 68 |
| | Thymalfasin | - | - |
| **Combination Therapies** | | | |
| | Azithromycin + chloroquine/ hydroxychloroquine | 6 | 40 |
| | Azithromycin + lopinavir/ritonavir | 5 | 29 |
| | Azithromycin + lopinavir/ritonavir + ribavirin | 5 | 35 |
| | Azithromycin + interferon + lopinavir/ritonavir + ribavirin | 2 | 11 |
| | Hydroxychloroquine + interferon + thymalfasin | 1 | 1 |
| | Hydroxychloroquine + lopinavir/ritonavir | 1 | 20 |
| | Interferon + thymalfasin | 5 | 16 |
| | Interferon + lopinavir/ritonavir | 2 | 8 |
| | Interferon + ribavirin | 1 | 1 |
| | Interferon + ribavirin + umifenovir | 1 | 1 |
| | Interferon + umifenovir + thymalfasin | 3 | 4 |
| | Lopinavir/ritonavir + ribavirin | 33 | 222 |
| | Lopinavir/ritonavir + umifenovir | 1 | 1 |
| | Lopinavir/ritonavir + interferon + ribavirin | 54 | 352 |
| | Ribavirin + umifenovir | 1 | 2 |
| | Umifenovir + thymalfasin | 1 | 2 |

**Table S3** Parameter estimates (relative standard errors, %RSE) from NLME model fit to available sample sites.$\beta$, rate target cells become infected; $\delta$, death rate of infected cells; $\gamma$, maximum rate of viral replication; $V_0$, the initial viral load. Urine and ocular $V_0$ were not estimable and fixed to LLOD/2 with $\gamma$ fixed to $e^1$

| | **Blood** | **Saliva/ Throat** | **Lower respiratory** | **Stool/ rectal** | **Urine** | **Breast milk** | **Ocular secretion** | **Nasal** |
|---|---|---|---|---|---|---|---|---|
| **Typical estimates** | | | | | | | | |
| $\boldsymbol{\beta}$ **[copies x day/ml]** | 0.0238 (29) | 7.25 x $10^{-4}$ (6) | 6.14 x $10^{-6}$ (6.5) | 1.37 x $10^{-5}$ (24) | 0.160 (15) | 2.07 (1838) | 4.70 x $10^{-3}$ (16) | 3.04 x $10^{-5}$ (19) |
| $\boldsymbol{\delta}$ **[1/day]** | 1.40 (230) | 0.588 (21) | 0.638 (46) | 0.572 (101) | 1.46 (110) | 1.56 (142) | 1.77 (30) | 0.796 (80) |
| $\boldsymbol{V_0}$ **[copies/ml]** | 1.84 (415) | 6.62 x $10^4$ (3) | 2.42 x $10^6$ (3) | 1.91 x $10^4$ (7) | 46.06* | 237.46 (358) | 42.52* | 66 x $10^4$ (2) |
| $\boldsymbol{\gamma}$ **[1/day]** | 1.76 (51) | 2.46 (11) | 6.36 (31) | 1.16 (38) | 2.72* | 1.27 (342) | 2.72* | 2.13 (257) |
| **Variance estimate %CV (%RSE)** | | | | | | | | |
| $\boldsymbol{\beta}$ | 232 (88) | 246 (44) | 439 (37) | 332 (35) | 67.7 | 415 (720) | 149 (394) | 352 (163) |
| $\boldsymbol{\delta}$ | 62.9 (122) | 87.5 (19) | 110 (35) | 87.0 (45) | 51.5 | 564 (884) | 47.2 (219) | 86.4 (49) |
| $\boldsymbol{V_0}$ | 483 (51) | 383 (12) | 282 (28) | 494 (29) | 570 | 510 (839) | 809 (195) | 443 (13) |
| $\boldsymbol{\gamma}$ | 10* | 10* | 10* | 0* | 10* | 10* | 10* | 10* |
| **Residual error** | 35 | 9.26 | 9.2 | 3.76 | 76.6 | 3.84 | 56 | 3.16 |
| **Residual error, nested** | 1.69 | 2.37 | 1.01 | 5.15 | | 4.11 | | 3.99 |

* fixed Parameters

**Table S4** Parameter estimates (relative standard errors, %RSE) from NLME model including covariate relationships. $\beta$, rate target cells become infected; $\delta$, death rate of infected cells; $\gamma$, maximum rate of viral replication; $V_0$, the initial viral load. Exponents of log-normal distributed parameters are reported. Drug effects can be interpreted as proportional change in δ.

| | **Parameter estimates (RSE%)** | **Bootstrap 95% CI** |
|---|---|---|
| **Typical estimates** | | |
| **$\beta$ [copies x day/ml]** | 1.02 x $10^{-3}$ (11) | 4.69x$10^{-6}$ – 0.078 |
| **$\delta$ [1/day]** | 0.60 (10) | 0.43 – 1.93 |
| **$V_0$ [copies/ml]** | 5.97 x $10^5$ (2) | 3.83 x $10^5$ – 1.14 x $10^6$ |
| **$\gamma$ [1/day]** | 2.29 (47) | 2.10 – 2.62 |
| **LRT on $\beta$ ++** | 9.4 x $10^{-4}$ (0.03) | 2.34 x $10^{-8}$ – 0.025 |
| **IFN on $\delta$ #** | 1.75 (0.06) | 1.05 – 2.66 |
| **LPVR/r on $\delta$ #** | 1.30 (0.07) | 0.75 – 1.86 |
| **Riba on $\delta$ #** | 1.47 (0.02) | 1.04 – 2.47 |
| **Age on $\delta$ +** | -0.07 (0.01) | -0.21 - -0.03 |
| **Sex on $\delta$ ++** | -0.15 (0.01) | -0.64 – 0.02 |
| **Variance estimate** | **%CV (RSE%) [shrinkage]** | |
| **$\beta$** | 437 (10) [41] | 231 – 568 |
| **$\delta$** | 60.2 (6) [24] | 42.0– 75.5 |
| **$V_0$** | 373 (6) [11] | 294 – 408 |
| **$\gamma$** | 10* | |
| **Residual error** | 1.16 (95) [6] | 0.02 – 18.9 |
| **Residual error, nested** | 7.95 (15) [6/2/0/2] | 5.09 – 10.12 |

* fixed Parameter
Covariate effects: # proportional effect, + power function, ++ absolute change

**Table S5** Some example sample sizes (number of patients per arm drug versus placebo) required to detect a significant difference in the proportion of undetectable viral loads after 7 days of treatment with the antivirals selected by the NLME model. Power was set at 90% for a 2-sided test and significance level of $p$<0.05.

| **Drug** | **Start Day 1** | **Start Day 3** | **Start Day 7** |
|---|---|---|---|
| **Ribavirin** | 185 | 225 | 317 |
| **Interferon** | 77 | 81 | 127 |
| **Interferon+ribavirin** | 23 | 27 | 41 |

#### Supplementary Figures

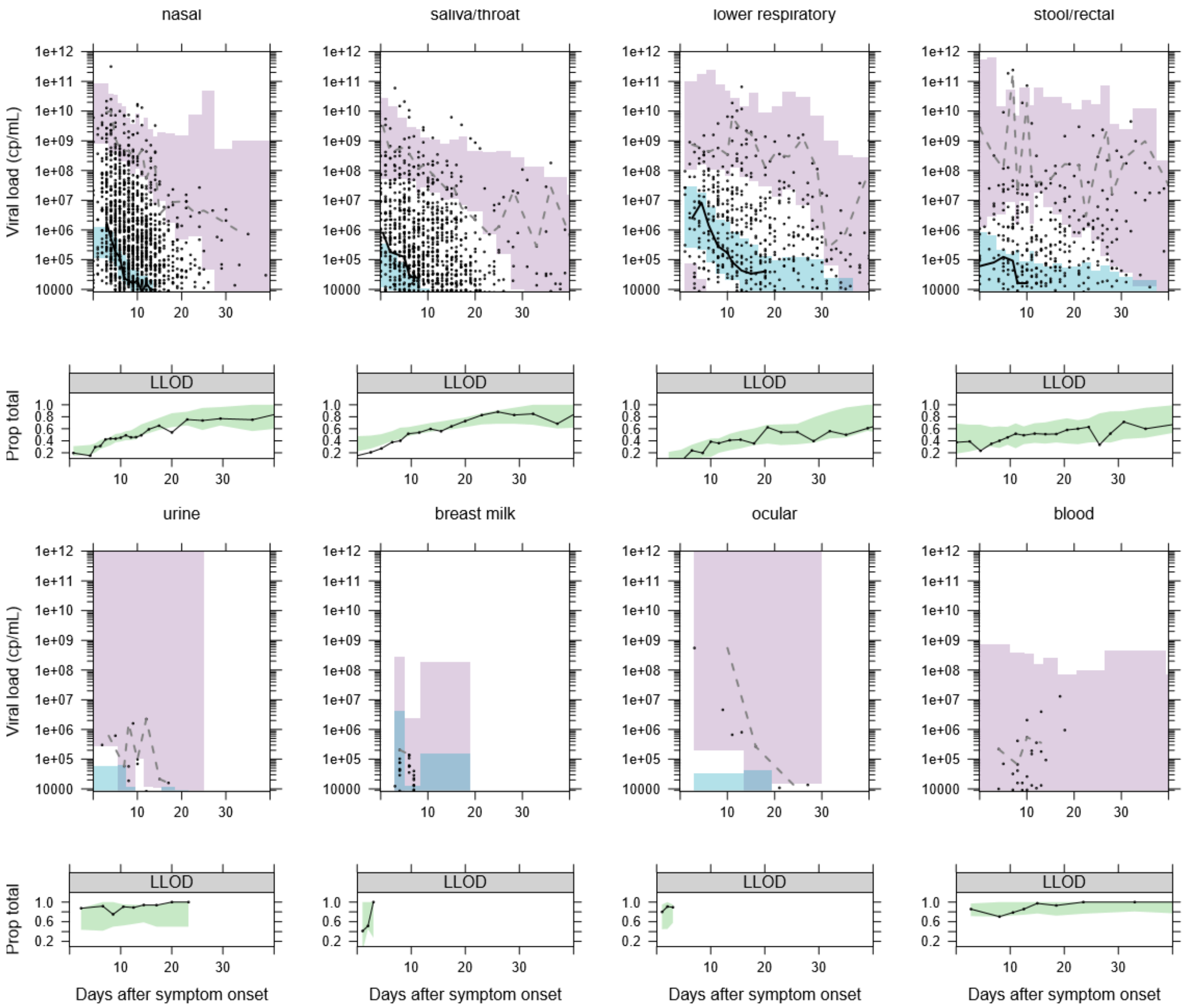

**Figure S1 Visual predictive checks for the NLME model fitted to viral load data to each sampling site**. For each site a plot of model simulations compared with observations is given for both the continuous data (upper) and the fraction of samples below the limit of detection (lower). Black circles are observed viral loads, purple shaded area is the 95% prediction interval of the simulated 2.5$^{th}$ and 97.5$^{th}$ percentile for comparison with the observed 2.5 and 97.5$^{th}$ percentile (dashed lines). The blue shaded area is the 95% prediction interval of the 50$^{th}$ percentile to compare with the continuous black line. In the lower plot the observed proportion of samples below the lower limit of detection (LLOD) are shown as a black line and compared with the 95% prediction interval of the model predicted proportion of samples below the LLOD (green shaded area)

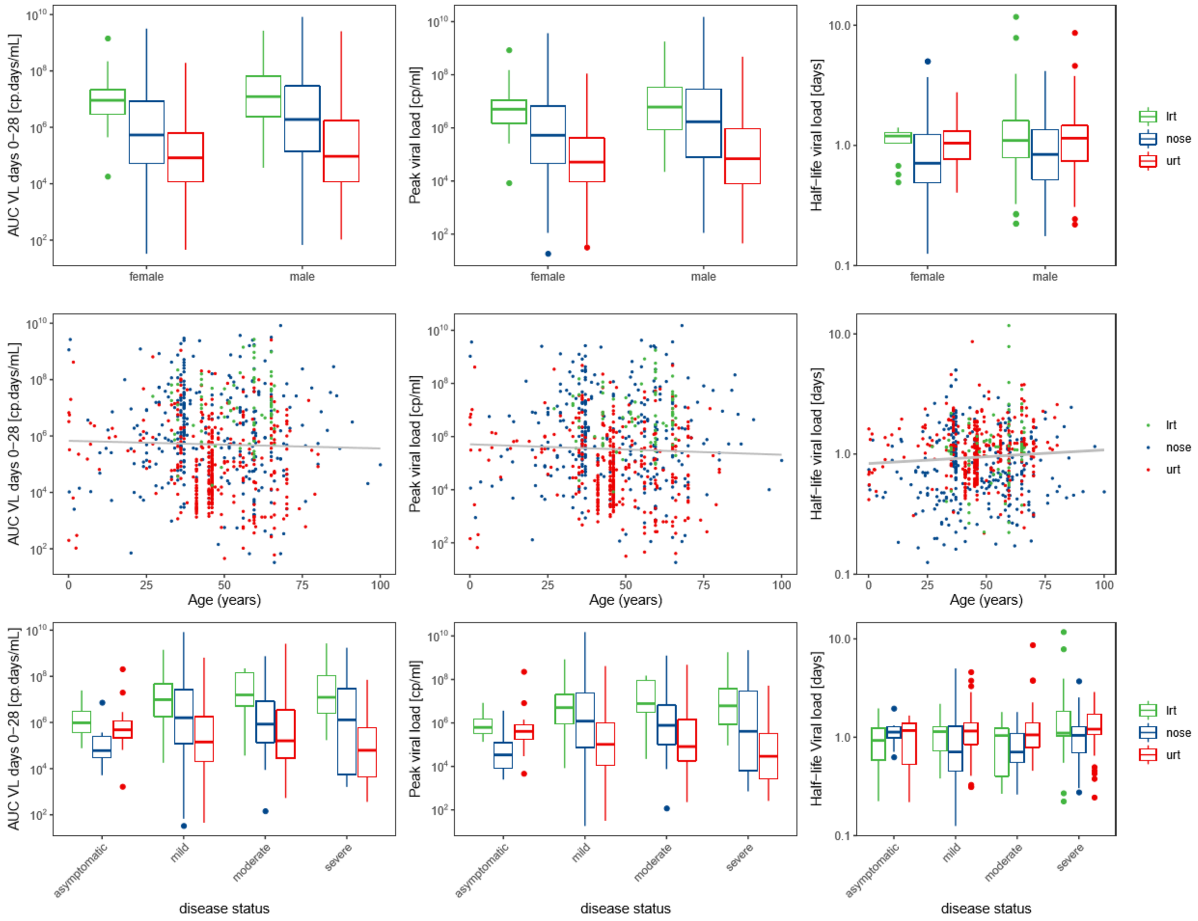

**Figure S2 NLME model derived parameters for respiratory sample sites.** Plot of model-predicted viral area under the curve from day 0-28 of symptom $AUC_{(0\text{-}28)}$, peak viral load and viral elimination half-life compared with sex, age and disease severity. lrt, lower respiratory tract; urt, upper respiratory tract

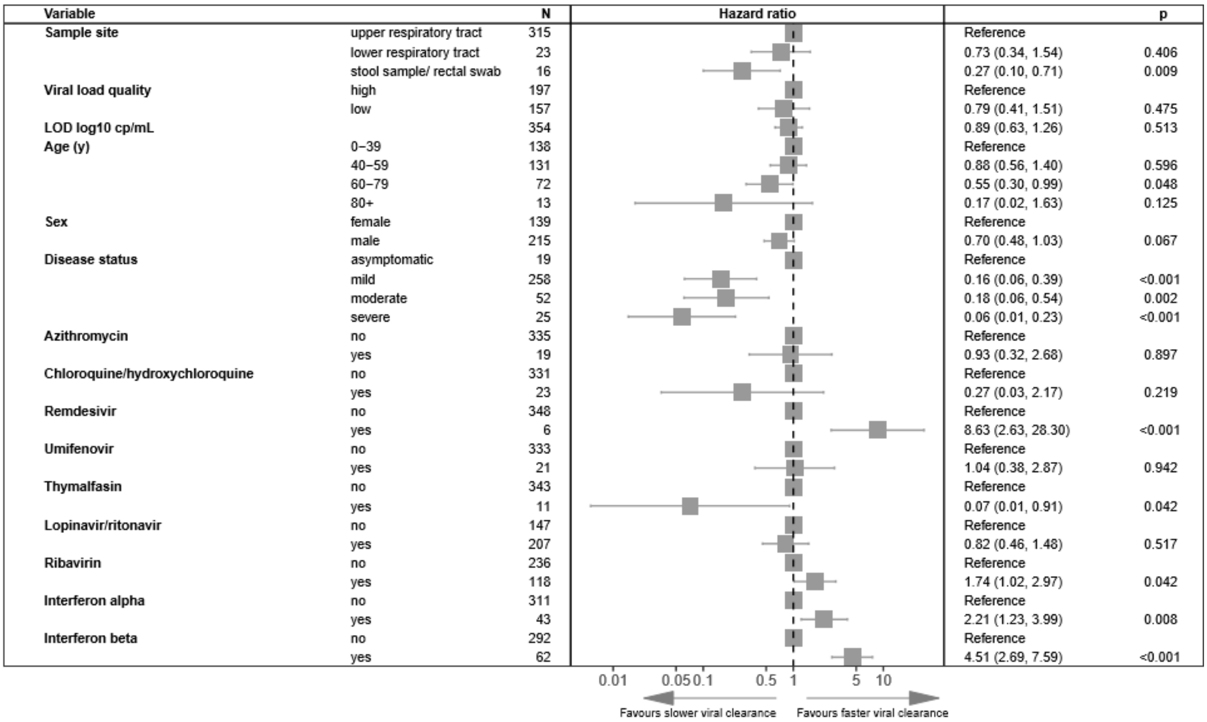

**Figure S3 Multivariable Cox proportional hazard results on all drug quality 1 and 2 data from respiratory and stool/rectal sampling sites with interferons split between alpha and beta.** Adjusted hazard ratios exceeding 1 indicate virus being more likely to become undetectable

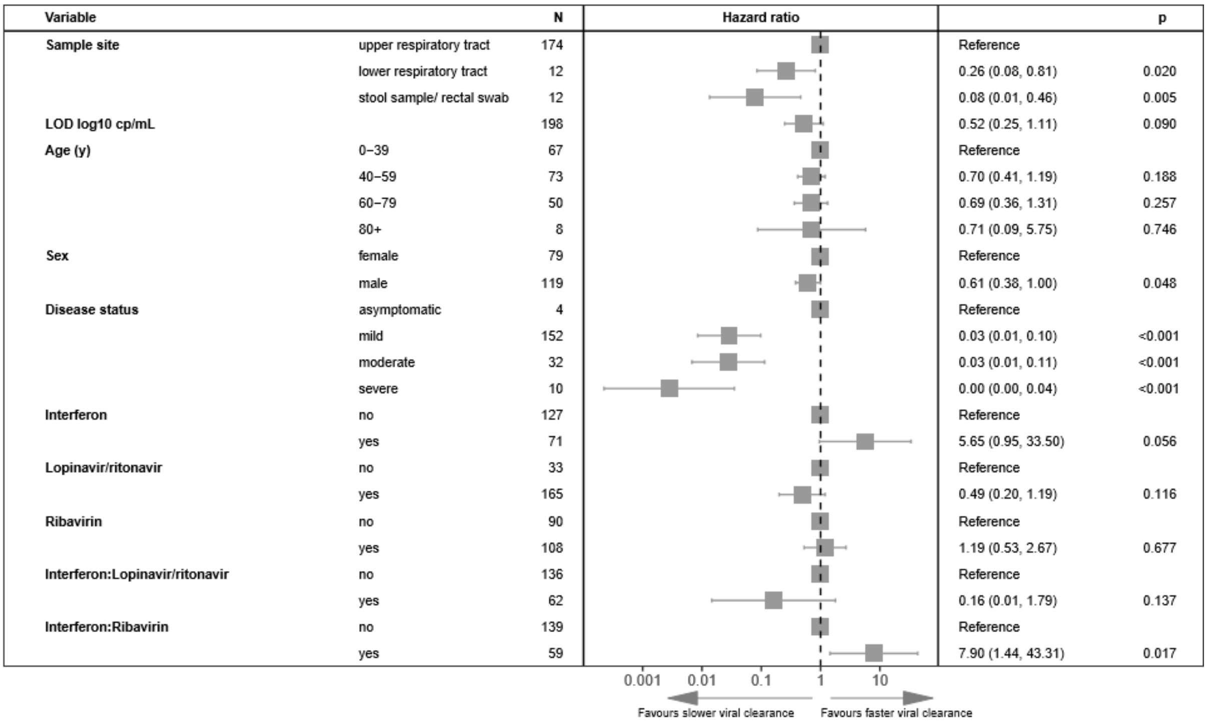

**Figure S4 Multivariable Cox proportional hazard results on viral load quality 1 data from respiratory sampling sites and stool/rectal sampling sites.** Adjusted hazard ratios exceeding 1 indicate virus being more likely to become undetectable

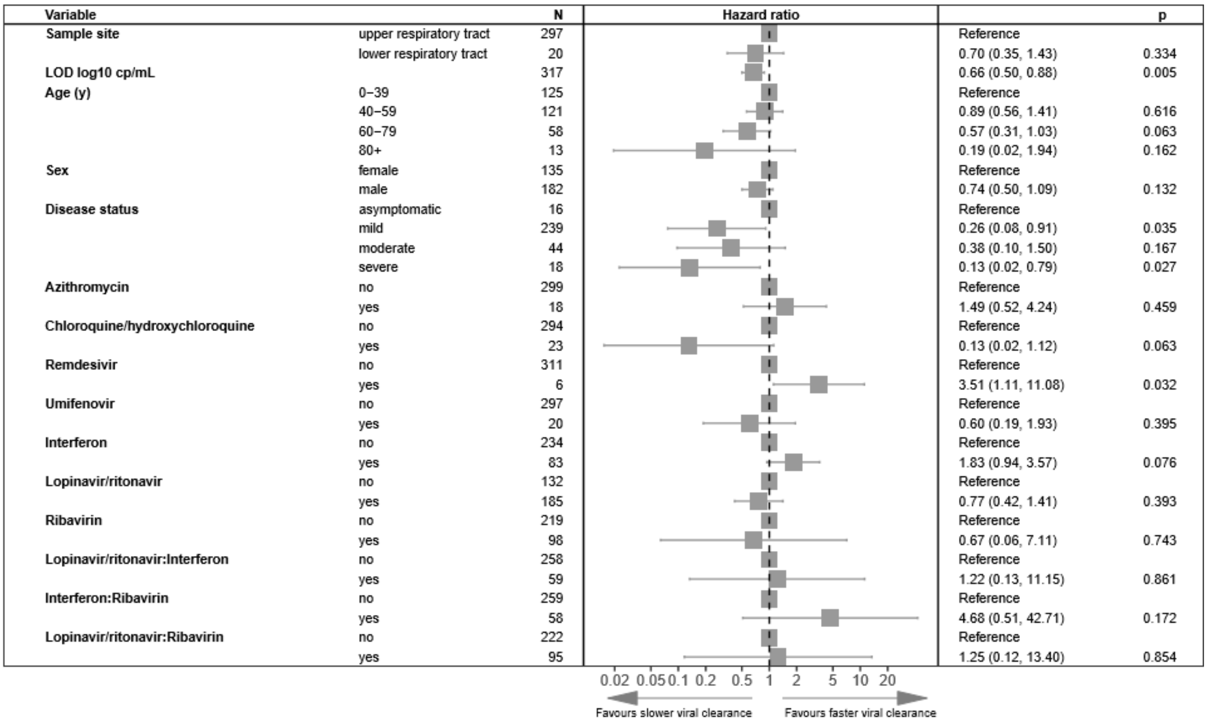

**Figure S5 Multivariable Cox proportional hazard results on viral load quality 1 data from respiratory sampling sites only.** Adjusted hazard ratios exceeding 1 indicate virus being more likely to become undetectable

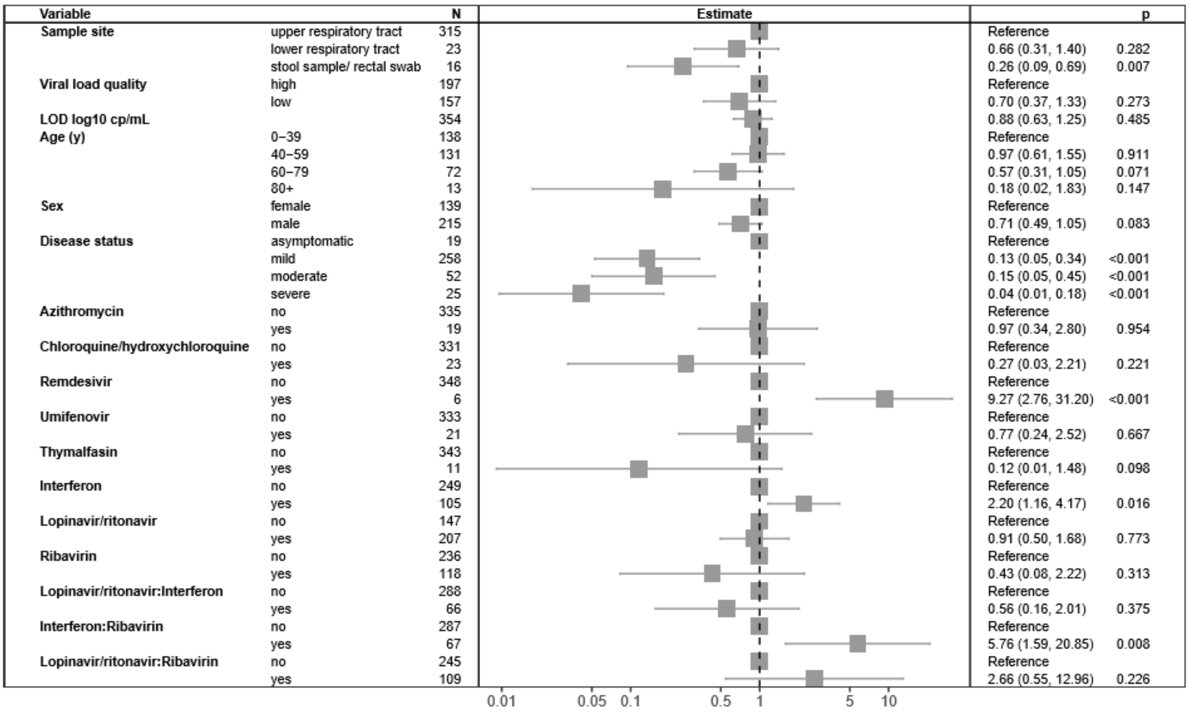

**Figure S6 Multivariable Cox proportional hazard results data excluding events with sampling frequencies over 3 days**. Adjusted hazard ratios exceeding 1 indicate virus being more likely to become undetectable

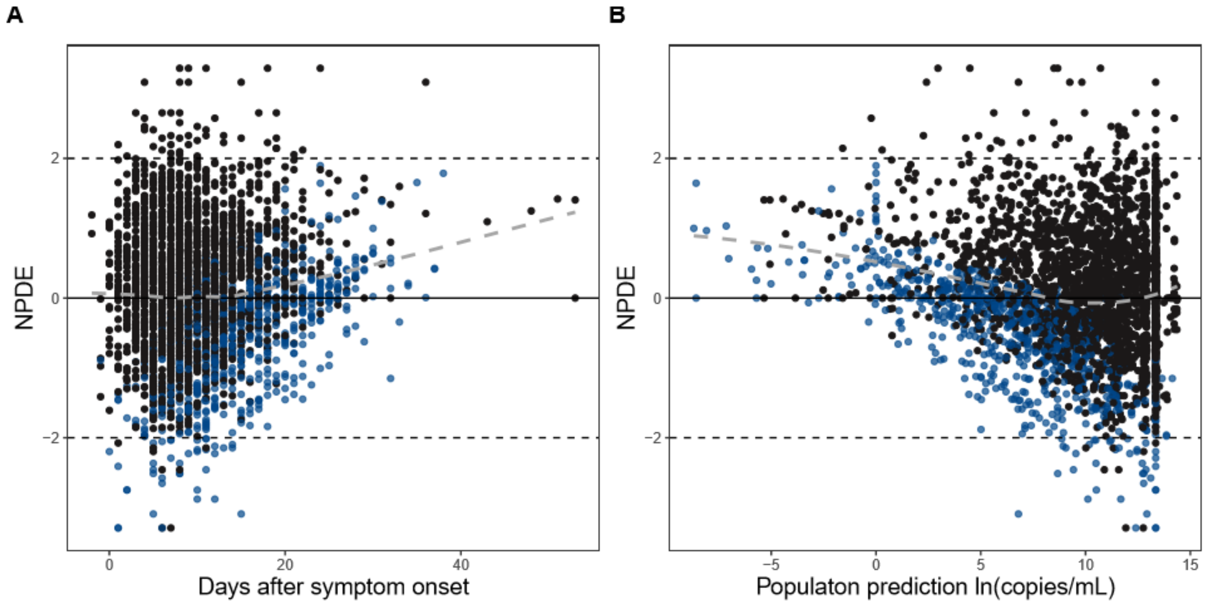

**Figure S7 Numerical prediction distribution errors (NPDE) based diagnostics for final model.** Black dots, data above lower limit of detection; blue dots, data below limit if detection

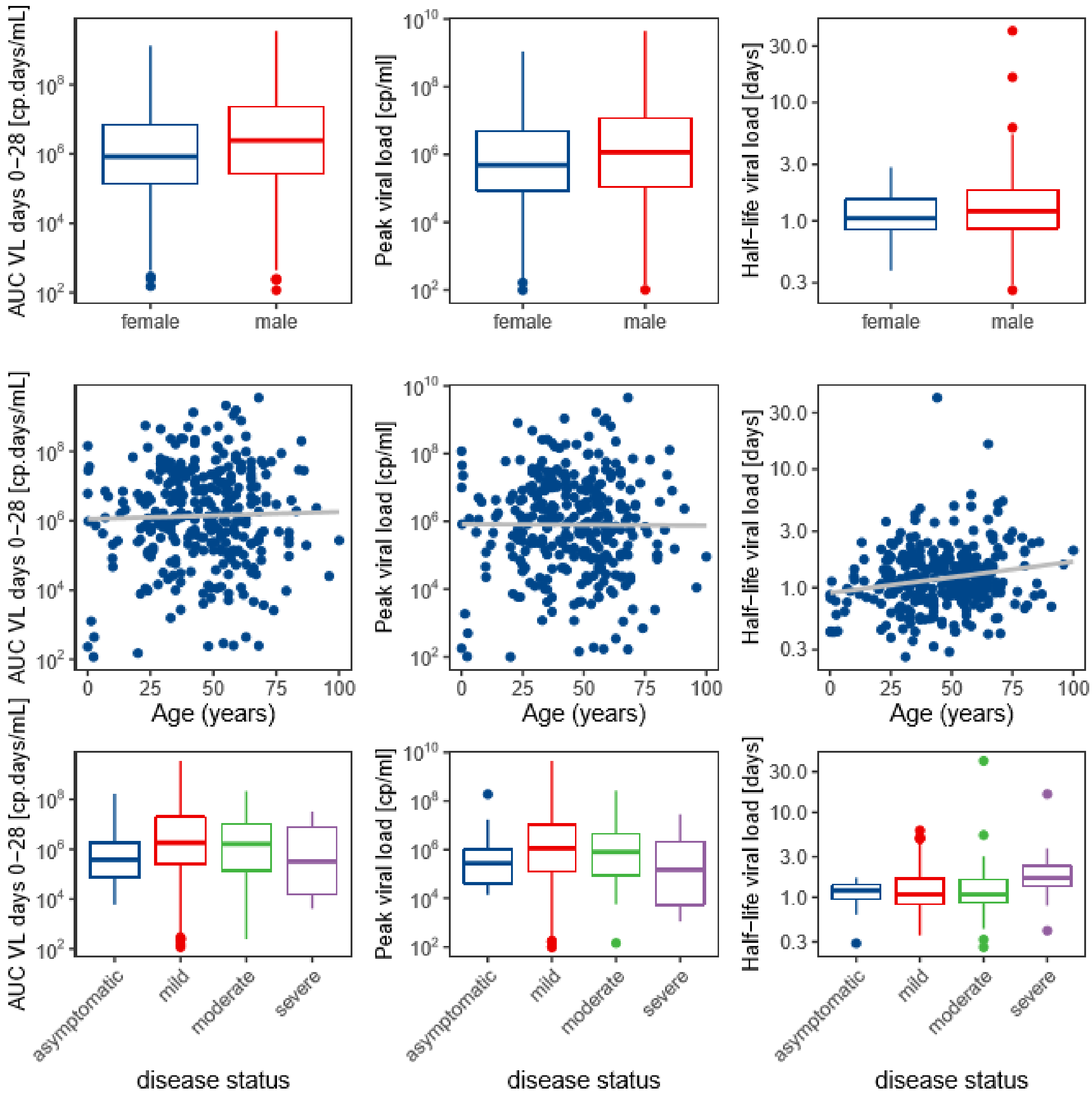

**Figure S8 NLME model derived parameters versus covariates for final model.** Plot of model-predicted viral area under the curve from day 0-28 of symptom onset; area under the viral load curve AUC(0-28), peak viral load and viral elimination half-life compared with sex, age and disease severity
